# Pulmonary sequelae at six months in children with SARS-CoV-2 infection: *A Single-Centre Study*

**DOI:** 10.1101/2023.03.10.23286644

**Authors:** Pothireddy Sharanya, Devendra Mishra, Anurag Agarwal, D Keerthana

**Author notes:** **Address for correspondence:** Dr. D Keerthana, Maulana Azad Medical College (University of Delhi) and associated Lok Nayak Hospital, 2, BSZ Marg, Delhi 110002, India.; E-mail id.

## Abstract

**Objective:** Pulmonary sequelae post SARS - CoV-2 infection have been reported in adults; however, there is scant literature regarding pulmonary dysfunction following SARS-CoV-2 infection in children. We studied the long term pulmonary sequelae in children who had SARS-CoV-2 infection.

**Methods:** This single center descriptive study conducted in a public sector tertiary care hospital in Northern India, from June, 2020 to October, 2021. We enrolled children aged 7-18 years admitted with SARS-CoV-2 infection and followed them up for 6 months. A detailed interval history was taken and pulmonary function tests were performed after 6 months, using a spirometer. A convenience sample of 40 children was enrolled. There were 21 males and the median (IQR) age was 13 (10.75, 17) years.

**Results:** Thirty percent of children (*n*=12) had pulmonary function abnormalities, which was of restrictive pattern in all. Children who were underweight had higher odds of developing pulmonary dysfunction following SARS-CoV-2 infection [OR (95% CI) 5.13 (1.19, 22.11); *P*=0.028]. There were no significant association with age, sex, severity of initial infection and oxygen requirement during the initial infection. Three children had persistence of dyspnea during follow up.

**Conclusion:** This study is the one of the first Indian studies regarding the pulmonary sequelae in children. A possibility of long term sequelae should be considered in children with history of SARS-CoV-2, presenting with suggestive complaints.

## Introduction

Novel coronavirus disease 2019 (COVID-19) caused by severe acute respiratory syndrome coronavirus 2 (SARS-CoV-2), has a lot of multiplicity in the spectrum of illness, but lung injury is responsible for most of the morbidity and mortality. Around 10% of patients are prone to respiratory failure or acute respiratory distress syndrome (ARDS).^1^ However, most of the children with COVID-19 are asymptomatic, and among the symptomatic children, many have only mild infection.^2^

Similar to severe acute respiratory syndrome (SARS), a histopathological progression has been seen in COVID-19 pneumonia, with intra-alveolar and interstitial fibrin deposition and chronic inflammatory infiltrates, after few weeks of diagnosis.^3^ Coronavirus infection can directly promote lung fibrosis. The nucleocapsid protein of SARS-CoV-1 is known to promote and enhance transforming growth factor-beta (TGF-β) signaling, which is a powerful pro-fibrotic signal.^4^ SARS-CoV-2 may also share such a feature.^5^

SARS-CoV-1 and Middle East Respiratory Syndrome (MERS), which are similar to SARS-CoV-2, are known to cause pulmonary sequelae, leading to pulmonary function impairment ranging from months to years.^6^ The long term effects on pulmonary function in COVID-19 survivors have not been studied in depth. Pediatric studies are very limited and no pediatric data is available from India yet. Therefore, we studied the occurrence of long term pulmonary sequelae in children with SARS-CoV-2 infection and the factors associated with it.

## Methods

This was a descriptive study, done in children aged between 7 to 18 years diagnosed with SARS-CoV-2 infection and admitted in the Department of Pediatrics of a tertiary care public hospital in Delhi from June, 2020 to October, 2021. Institutional Ethics Committee of our institute cleared the study. The inclusion criterion was children who were positive for SARS-CoV-2 infection, either by reverse transcriptase-polymerase chain reaction (RT-PCR) or rapid antigen test, and admitted in the department of pediatrics. Those with any of the following features were excluded: diagnosed with chronic respiratory diseases like cystic fibrosis, interstitial lung diseases and asthma; pre-existing neuromuscular disorders like muscle dystrophies and spinal cord injury that may affect pulmonary function tests; pre-existing conditions that may affect pulmonary function test like lung surgery, known sequelae of pulmonary tuberculosis, kyphosis, scoliosis and chest wall deformities; neurodevelopmental disorders, where poor cognitive ability may hinder performing the pulmonary function test; and either not residing in Delhi-NCR or unwilling to return for follow up. Additionally, those who were having any acute illness at the time of pulmonary function test were asked to revisit after its resolution within two weeks period. The primary outcome variable was measures of lung function on pulmonary function test viz., forced vital capacity (FVC), forced expiratory volume in first second (FEV1), and ratio of FEV1 and FVC.

The reported proportion of adult patients with SARS-CoV-2 infection having pulmonary sequelae is 25.5%.^7^ The calculated sample size for similar sample proportion with 10% margin of error and 90% confidence is 52. We planned a sample size of convenience of at least 40 children.

The eligible subjects were identified by two methods viz., screening the hospital records of patients admitted from June, 2020 to December, 2020, and prospectively from inpatients with COVID-19 admitted from January, 2021 to October, 2021. The case files of the eligible subjects admitted due to SARS-CoV-2 infection from June, 2020 to December, 2020 were identified from the records. They were screened and contact details were noted. Their parents were contacted telephonically and were informed about the study, and requested to follow up after 6 months of discharge, if interested in enrolling. At the follow up visit, subjects who satisfied the inclusion and exclusion criteria were enrolled into the study, after informed written consent of parents, and assent from children more than 7 years of age. The symptoms at presentation, baseline investigations if any, duration of hospital stay and course during hospital stay were noted from the hospital records.

Eligible subjects were also prospectively identified from January, 2021 to October, 2021, from the inpatients admitted due to SARS-CoV-2 infection in the hospital. A detailed clinical examination was done and children who satisfied the inclusion and exclusion criteria were enrolled into study after informed consent and assent. Relevant demographic and clinical details were recorded. These families were also requested to follow up after 6 months of discharge. All eligible subjects were telephonically informed about the date of follow up visit, one month prior to the visit, and again within one week of follow up date, if they missed the visit.

A structured form was used to collect the details of the patients. It contained sections regarding the demographic data, and presence and duration of various symptoms like fever, cough, coryza, sore throat, vomiting, abdominal pain, diarrhea, body pain, breathing difficulty, tachycardia and seizures. Results of laboratory investigations including hemogram, D-dimer, C-reactive protein (CRP), interleukin-6 (IL-6), procalcitonin (PCT), ferritin, blood urea, serum creatinine, liver function test, prothrombin time, international normalized ratio (INR) and chest X-ray reports, if done were collected. Duration of hospital stay, oxygen requirement, treatment received, and outcome were also recorded.

At the follow up visit, all the participants were asked about the resolution of symptoms and current issues if any. General physical examination and systemic examination were done. Anthropometry which includes weight, height and body mass index (BMI) were noted and standard deviations were calculated using WHO charts for height and weight and centiles for BMI.^8^

Subjects who were unable to perform the maneuvers of pulmonary function test after five attempts were excluded from the study. Pulmonary function test was carried out according to the spirometry guidelines given by American Thoracic Society/ European Respiratory Society,^9, 10^ and Schiller SP-260 PC spirometer (Schiller Healthcare India Pvt. Ltd. Delhi) was used. During the visit, subjects were asked to come after having a light breakfast. A single trained respiratory technician performed all the tests. Children were demonstrated the technique of spirometry, and were made to rest for five to ten minutes before carrying on the test. They were asked to take 2–3 tidal breaths in and out and to inhale deeply with the lips sealed tightly around the mouthpiece. They were then asked to blow air through the mouthpiece as fast as possible (to blast air from the lungs) and to continue until no air is left to exhale. The test was repeated at least three times for consistent results. The obtained results were assessed for acceptability and repeatability.^10^ The measured values were recorded and compared with predicted values. Best of the three attempts were taken into consideration. After the test, child was subjected to 6 minutes’ walk test and pulmonary function test was repeated immediately. Predicted values of FVC, FEV1/FVC, and FEV1 given by Knudson,^11^ based on age, sex and height were used in the software to calculate the percentage of predicted values. FVC<80% of predicted, FEV1<80% of predicted, FEV1/FVC<90% in below 11 years and <75% above 12 years of age was considered abnormal.^12^

### Data analysis

Data were entered in microsoft excel sheet by a single researcher, and tested for missing values. Categorical data (demographic and clinical features) are presented as proportions and continuous variables (age, sex, anthropometry, PFT values) are presented as median (IQR). All analyses were done using Epi Info software. The risk of pulmonary dysfunction was compared for patients with different risk factors by univariate analysis using odds ratio (OR) with 95% confidence interval. A *P* value <0.05 was considered statistically significant.

## Results

The study enrolled 40 patients after pulmonary function testing on follow up of 6 months. The baseline characteristics of the study subjects are shown in Table I. The median age of the study cohort was 13 (10.75, 17) years. The median (IQR) weight and height *z*-scores were -0.25 (−1.0, +0.33) and - 0.43 (−0.79, -0.4), respectively. Based on the BMI, 30% of children (*n* =12) were underweight (BMI<5%) and 8% of children (*n*=3) were obese (BMI >95%). According to Government of India (GOI) staging of severity of COVID-19, the participants were divided into four groups. Majority of the children had mild disease (50%). Among the symptomatic children (*n=35)* the most common symptom found was fever (77%), vomiting (32.5%), headache (28%). Only six children (15%) admitted required oxygen support. Four of these were classified as severe, and two others were with moderate disease who required venturi and nasal prong support.

Blood investigations and imaging were not done in all patients as per the guidelines in force during the pandemic, and investigations were done based on disease severity. Majority of children had normal leucocyte counts. Thrombocytopenia was found in 34% children. Serum ferritin was elevated in 75% of children (*n*=6) and procalcitonin was elevated in all these children (*n*=6). Majority of children had normal chest *x*-ray findings.

Of the 40 children evaluated, 30% had FVC<80%, 25% had FEV1<80%. All children had normal FEV1/FVC ratio. All of the children with abnormal parameters had restrictive ventilatory defect. After 6 minutes’ walk, there was no significant change compared to initial PFT parameters, except in one child who had fall in FEV1<80% (Table II).

Pulmonary dysfunction was present in 33% of males and 31% of females. There was no statistically significant difference in the age, gender, severity, *x*-ray findings and pulmonary dysfunction in children (Table III). About 58% of underweight children had respiratory dysfunction on follow up. The odds of long term sequelae were higher for underweight children as compared to normal weight children [OR (95% CI) 5.13 (1.19, 22.11); *P*=0.028]. The asymptomatic children did not have abnormal spirometry, whereas 50% (*n*=2) children in severe group had pulmonary dysfunction. Of the 14 children with normal chest *x*-ray, 10 children (71%) had dysfunction. Two out of six patients who did not require oxygen therapy during the acute stage, were found to have pulmonary sequelae in spirometry.

## Discussion

In this single-center descriptive study, we found 30% of children with SARS-CoV-2 infection to have pulmonary dysfunction in the form of abnormal spirometry parameters at six months after the infection. Being underweight was the only factor significantly associated with the risk of long term pulmonary sequelae. One of the earliest studies evaluating pulmonary dysfunction following respiratory virus infection, reported that 16% of the adult patients were affected after six months of the 2003 SARS epidemic.^13^ Zhao, et al.^7^ reported that lung function abnormalities were seen in around 25% of the 55 adult patients followed up after three months of SARS-CoV-2 infection.

Very few pediatric studies have addressed this question. A recent study by Palacios, et al.^14^ followed up children for 6 months following infection. Pulmonary function abnormalities were seen in 23% and 10%, at 3 and 6 months of follow up, respectively. A study from the US reported spirometry abnormalities in 10% of the subjects.^15^ Relatively lesser prevalence of respiratory dysfunction in comparison to our study could be because outpatients with mild disease and competitive athletes comprised the majority of their study cohort. No child with pulmonary dysfunction on follow up was seen in the studies by Chiaria, et al. (*n*=61) and Bottinio, et al. (*n*=16), possibly because both these studies enrolled only mildly symptomatic and asymptomatic children.^16, 17^

The mechanism behind post covid lung injury is still a matter of debate. A prolonged pro inflammatory response related to SARS-CoV-2 infection can provoke an atypical response of the immune system and mast cells, promoting a cascade of events affecting the respiratory and immune systems.^18^ SARS-CoV-2 can induce pulmonary fibrosis by promoting the upregulation of pro-fibrotic signaling molecules, including transforming growth factor-beta (TGF-β). The diffuse alveolar damage, airway inflammation and intra alveolar thrombosis progress to late stages of lung repair involving lung fibrosis and interstitial remodelling.^19^

The pulmonary dysfunction was restrictive type in all patients, which is similar to adult studies.^20, 21^ Majority of the children enrolled were asthmatics in the pediatric studies which could be the reason for obstructive pattern predominating on follow up.^14, 15^

We found a significant association of being underweight (BMI <5^th^ centile) with pulmonary dysfunction. Underweight children are more prone to infections per se, and they are also at increased risk of secondary bacterial infections. Association of both underweight and obesity with severity of COVID-19 disease has previously been reported in adults.^22^ However, no such association has been seen in pediatric patients.^14-17^

The occurrence of long term sequelae was not significantly associated with severity of initial infection. Similar results have been elucidated by Vezir, et al.^23^ where spirometry values of 57 patients showed no significant difference between the asymptomatic/ mild and moderate/ severe groups. A German study has also quoted that persistence of respiratory symptoms is not related to the severity of infection.^24^ Three patients had symptoms on follow up, and all of them reported exertional dyspnea. Recent pediatric studies have reported that shortness of breath, cough, exercise intolerance as the common symptoms noted in their follow up cohort.^25, 26^

Our study had many limitations, especially the small sample size. This was due to designation of the institute as an exclusive COVID-facility, and even the recovered patients were unwilling to come for follow up to the institute. Additionally, the baseline lung function parameters of the participants in the study were not available. This could be a confounding factor; even though, history of prior respiratory symptoms was taken at the follow up to rule out this possibility. More sensitive investigations like plethysmography and gas exchange could have been done for evaluation of the pulmonary function, though these are meant for research settings and not routinely available for clinical use.

We found that pulmonary dysfunction was present in children affected by SARS-CoV-2, and children with underweight are more prone to pulmonary dysfunction on follow up. Our study has exemplified that despite milder presentation in children than adults, children may suffer long term sequelae secondary to covid infection. Physicians may need to actively rule out the possibility of pulmonary dysfunction in SARS-CoV-2 affected children, who present with suggestive complaints.

## Data Availability

All data produced in the present study are available upon reasonable request to the authors.

## Contributors

DM conceptualized and planned the study, did the statistical analysis, and finalized the manuscript. PS assisted in planning the study and statistical analysis. AA provided intellectual inputs regarding conduct of the study and manuscript preparation. DK assisted in planning the study, outcome assessment and manuscript preparation. Both PS and DM prepared the initial draft of the manuscript. All authors approved the final manuscript, and agreed to be accountable for all aspects of the study.

## Acknowledgement

Department of Pulmonary Medicine, Lok Nayak Hospital, Delhi for help in the conduct of pulmonary function tests.

---

**Figure 1.**
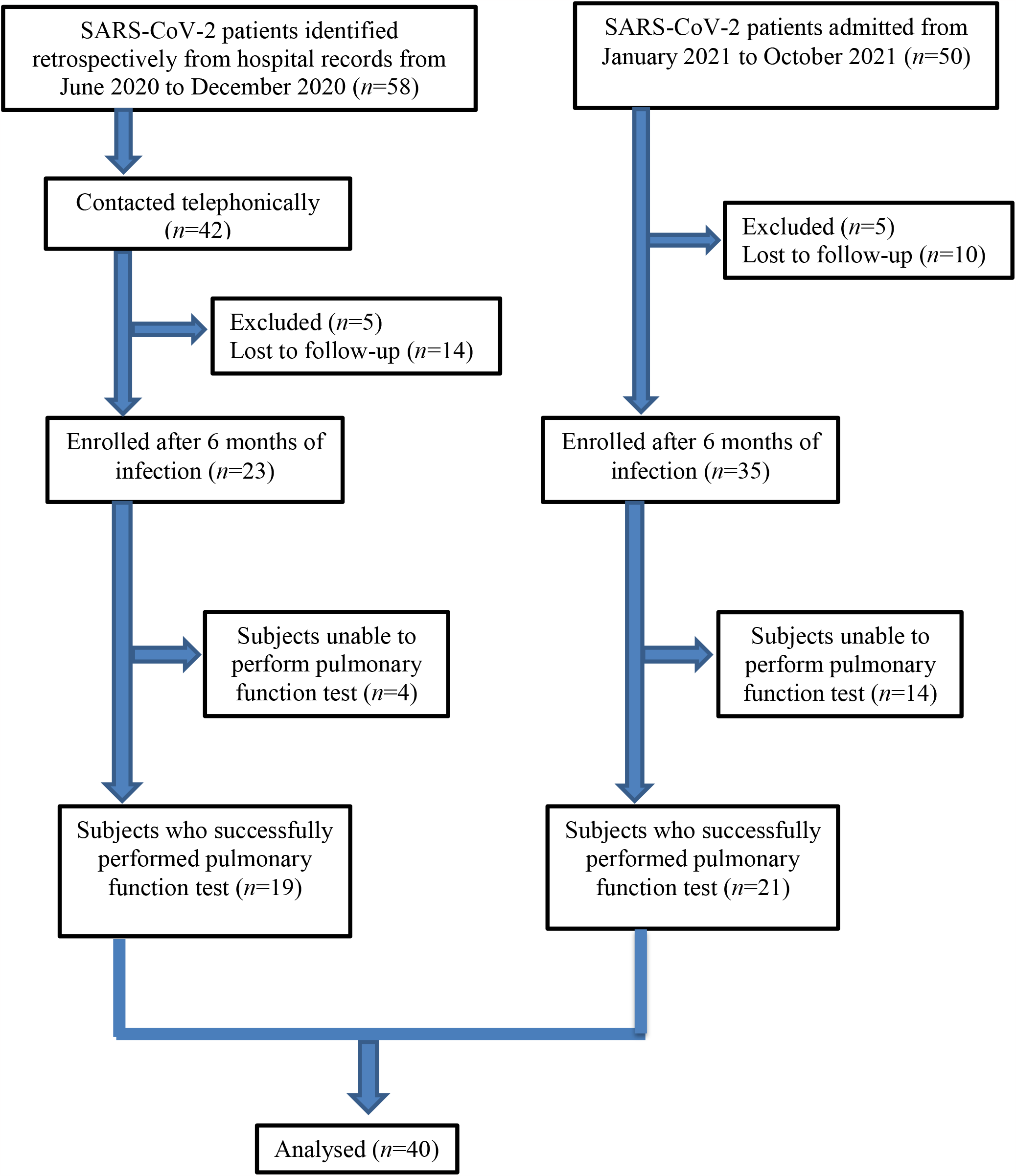
Study flow chart.

